# Preserved white matter structure at the grey matter–white matter interface despite widespread cortical thinning in early psychosis

**DOI:** 10.1101/2025.05.13.25327498

**Authors:** Yoshito Saito, Warda T. Syeda, Yasmin Gurleyen, Marta Rapado-Castro, Maria A. Di Biase, Cassandra M.J. Wannan, Remika Mito, Christos Pantelis

## Abstract

**Background:** Early psychosis is characterized by widespread cortical grey matter (GM) reductions. White matter (WM) alterations have also been reported. The GM–WM interface may represent a link between cortical GM and WM alterations that remains understudied. As the interface matures later in development than deep WM, it may be particularly vulnerable to neurodevelopmental disruptions in schizophrenia-spectrum disorders. However, its complex fiber architecture and proximity to the cortex pose challenges for conventional diffusion tensor imaging.

**Methods:** We applied an advanced diffusion MRI approach to assess fiber density (FD) in the GM–WM interface of 78 individuals with early psychosis (age 22.0 ± 3.0, 29.5% female) and 78 controls (age 21.8 ± 3.2, 29.5% female). We examined group differences in FD at the GM–WM interface and cortical measures, and assessed spatial correlations between regional effect sizes of FD and cortical measures.

**Results:** Widespread cortical thinning was observed, whereas no significant group differences were identified in FD at the GM–WM interface. This finding remained robust after accounting for partial-volume effects and was consistent with the template-space analysis. The spatial distribution of FD effect sizes showed significant positive correlations with those of cortical thickness and surface area.

**Conclusions:** Despite widespread cortical GM alterations, WM microstructure at the GM–WM interface was relatively preserved in early psychosis, while spatial correspondence between GM and WM measures at the interface was observed. These findings suggest that cortical alterations are pronounced in early psychosis, whereas WM changes may manifest later in the course of illness.

## INTRODUCTION

Schizophrenia-spectrum disorders are psychiatric conditions characterized by positive and negative symptoms and cognitive dysfunction (1). Neuroimaging studies of schizophrenia-spectrum disorders have consistently demonstrated widespread grey matter (GM) reductions alongside white matter (WM) alterations (2–5). These widespread alterations in cortical GM have also been consistently observed in early psychosis, while findings in deep WM are heterogeneous in their spatial distribution and effect sizes across studies (6–12). GM alterations have been suggested to propagate along WM connections and precede observable WM abnormalities in schizophrenia (13–15). However, how cortical pathology is directly associated with connected WM remains unclear. Examining the association between cortical GM and the directly adjacent WM at the GM–WM interface, which represents the axonal entry and exit zone of cortical neurons, may clarify this relationship. While our previous work found a negative correlation between cortical thickness and adjacent WM integrity measured by fractional anisotropy (FA), this study focused on WM regions 4–6 mm beneath the GM–WM interface, leaving the interface itself understudied (16). Further, the GM–WM interface is also important from a neurodevelopmental perspective, as it shows a later pattern of myelination during development compared with long-range axons, which potentially makes the interface more vulnerable to compromise in neurodevelopmental conditions (17).

Post-mortem brain studies suggest that the most superficial layers of WM are implicated in schizophrenia-spectrum disorders, demonstrating microstructural changes, including reduced myelinated fiber volume and increased interstitial WM neurons (18–22). However, while several neuroimaging studies have demonstrated alterations in the superficial white matter adjacent to cortical GM in frontal regions (16,23–25), the GM–WM interface has not been investigated directly. This may be due to several features that make the GM–WM interface difficult to characterize using conventional diffusion tensor imaging (DTI), the most commonly adopted method for studying WM (Figure 1). First, the GM–WM interface is susceptible to partial volume effects from GM, which can confound diffusion measurements (16). Second, the GM–WM interface contains multiple crossing fibers due to the intersection of short-range U-fibers and long-range association fibers, which cannot be resolved by the single-tensor model used in DTI (26–28). The presence of crossing fiber populations and partial voluming of tissue types makes DTI-based measurements (e.g., FA) highly non-specific at the interface, and problematic for interpreting differences in these metrics as representing changes to “WM integrity” (29).

**Figure 1.**
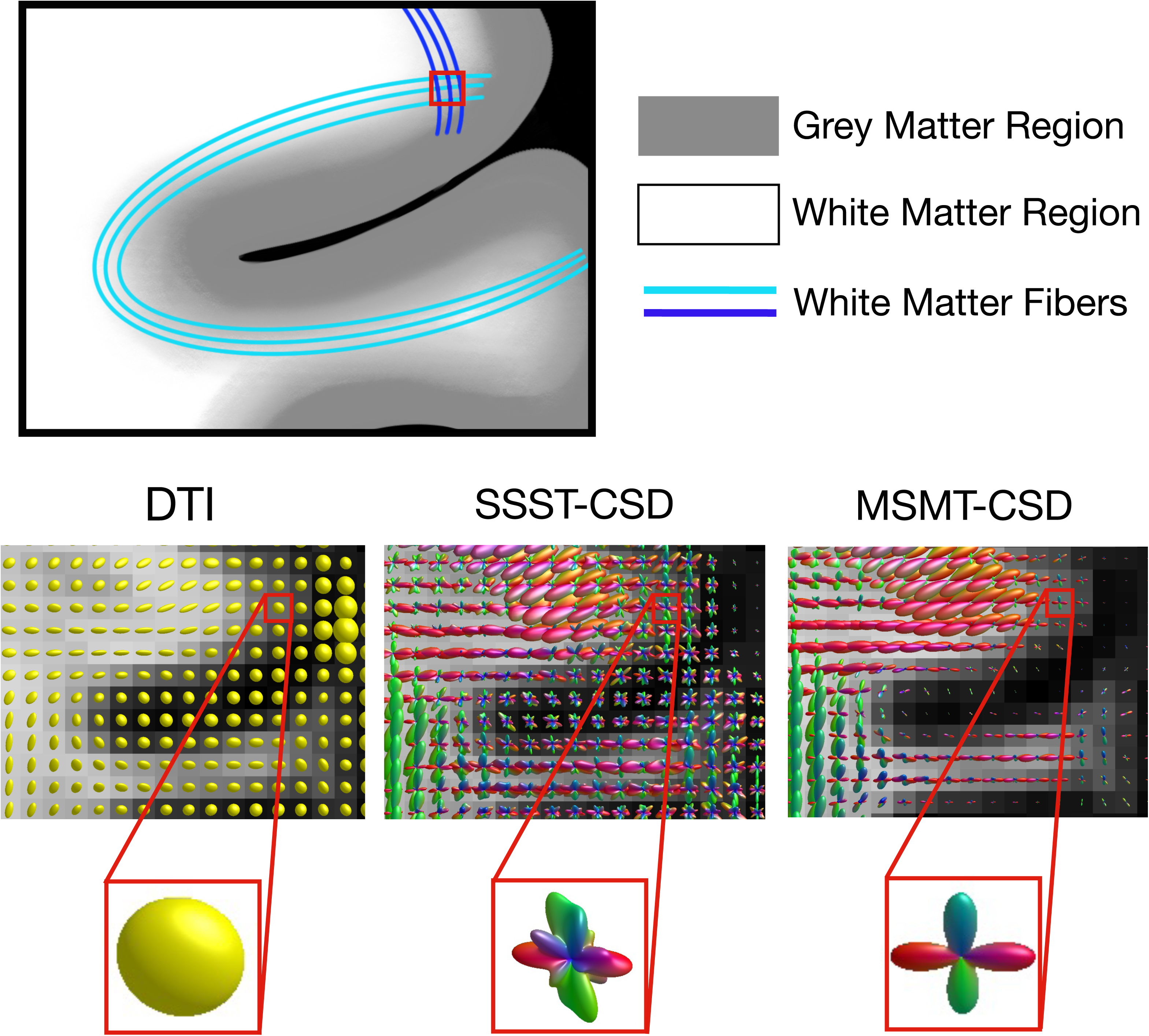
Differences in modeling white matter fibers at the grey matter–white matter (GM–WM) interface using diffusion tensor imaging (DTI), single-shell single-tissue constrained spherical deconvolution (SSST-CSD), and multi-shell multi-tissue CSD (MSMT-CSD). This figure illustrates the differences in white matter fiber representations at the GM–WM interface across three approaches: DTI (left), SSST-CSD (middle), and MSMT-CSD (right). As the schematic diagram (top) shows, the GM–WM interface contains complex fiber populations, including short-range cortico-cortical connections (often referred to as U-fibers) and long-range connections. The interface is therefore complex in architecture and may contain a high number of crossing-fiber populations (for example, in the highlighted voxel (red square)). CSD is able to model crossing fiber populations, whereas the DTI model fails to resolve crossing-fiber structures. Measures like fractional anisotropy are averaged across the whole voxel, meaning changes in these measures can be difficult to interpret in areas with crossing fiber populations. Furthermore, the signal estimated by SSST-CSD at the GM–WM interface is influenced by the grey matter compartment; however, by using MSMT-CSD, we can ensure that only the white matter fraction is modeled, while accounting for grey matter and CSF in areas affected by partial voluming. The fiber density (FD) measure used in this study is therefore specific to white matter and can be extracted for each distinct fiber population (i.e., each lobe modeled by the fiber orientation distribution (FOD) functions shown for each voxel).

To address these limitations, we implemented constrained spherical deconvolution (CSD), which enables modeling of crossing fiber populations. In particular, by using a multi-shell multi-tissue CSD (MSMT-CSD) approach, we minimized the influence of non-WM signals, enabling more specific analysis of WM within the GM–WM interface (30). Using this advanced modeling approach, we characterized fiber density (FD) (31,32) at each specific WM ‘fixel’ (fiber population within a voxel), thereby overcoming limitations of DTI-based measures by providing a more specific measure that characterizes WM at the GM–WM interface.

Using this advanced diffusion MRI approach, the present study aimed to (1) compare the WM structure at the GM–WM interface between individuals with early psychosis and healthy controls, and (2) examine the relationship between the GM–WM interface and overlying cortical morphology. We hypothesized that (1) individuals with early psychosis would exhibit altered fiber density at the GM–WM interface compared with healthy controls in frontal regions; and (2) fiber density at the interface would show a negative association with cortical alterations.

## METHODS AND MATERIALS

### Participants

This study included 78 individuals with early psychosis from the Human Connectome Project for Early Psychosis (HCP-EP) and 78 age- and sex-matched healthy controls from two datasets: HCP-EP and the Human Connectome Project in Development (HCP-D) (33,34). While the HCP-EP dataset includes both individuals with early psychosis and healthy controls, given the difference in group size, we used the HCP-D dataset to increase the number of control participants. Among the early psychosis individuals, we included those diagnosed with non-affective schizophrenia-spectrum disorders in the DSM-5 within five years of their first psychotic episode. In addition to the exclusion criteria for the HCP-EP and HCP-D datasets, healthy controls with a first-degree family history of schizophrenia were also excluded. Propensity score matching was used to select healthy controls from both datasets, ensuring close matching in terms of group size, age, and sex (35,36). Details on the datasets and matching procedures are provided in the Supplementary Material.

### Clinical assessment

The severity of clinical symptoms was assessed by the Positive and Negative Syndrome Scale (PANSS) (37).

### MRI acquisition and preprocessing

T1- and diffusion-weighted images were obtained from the HCP-EP and HCP-D studies within the HCP consortium. MRI data were acquired using 3T Siemens MAGNETOM Prisma scanners at Indiana University, Brigham and Women’s Hospital, McLean Hospital, and Harvard University. Both studies followed equivalent imaging protocols, including T1-weighted images with 0.8 mm isotropic resolution and multi-shell diffusion-weighted images (1.5 mm isotropic, TR = 3230 ms, TE = 89.20 ms) with two b-values (1500 and 3000 s/mm²), and 92 diffusion directions per shell and 14 b0 images (all shells acquired in both AP and PA directions). Further sequence details are available in the original protocol publications (34,38).

Minimally preprocessed T1-weighted images were made available by the HCP consortium. Cortical thickness and surface area were extracted based on Destrieux atlases using FreeSurfer (version 6.0.0) (https://surfer.nmr.mgh.harvard.edu) (39).

Raw diffusion-weighted images were preprocessed locally with the HCP minimal preprocessing pipeline, including EPI distortion, eddy current, and motion correction with FSL (FMRIB’s Software Library, www.fmrib.ox.ac.uk/fsl) (version 6.0.4) (40). The quality of preprocessed images was assessed with two quality control frameworks of FSL (version 6.0.4), Quality Assessment for dMRI (QUAD) and study-wise QUAD (SQUAD) for diffusion-weighted images, including automated subject-level and study-level reports providing representative slice images and summarizing motion, outlier slices, and signal quality metrics (41).

### Assessment of GM–WM interface

To assess WM changes within the GM–WM interface, we quantified fiber density (FD), which is commonly used within the ‘Fixel-based analysis’ (FBA) framework – a statistical analysis framework for estimating fiber-specific properties at the sub-voxel level (Figure 2) (32,42,43). FD is based on modeling specific <u>fi</u>ber populations within a vo<u>xel</u> (known as ‘fixels’), and is approximately proportional to total intra-axonal volume under certain conditions (31,44). The FD value was obtained by modeling multi-shell DWI data with multi-shell multi-tissue constrained spherical deconvolution (MSMT-CSD), which separates anisotropic WM signal from isotropic GM and CSF, and mitigates partial-volume effects prior to FD computation (30).

**Figure 2.**
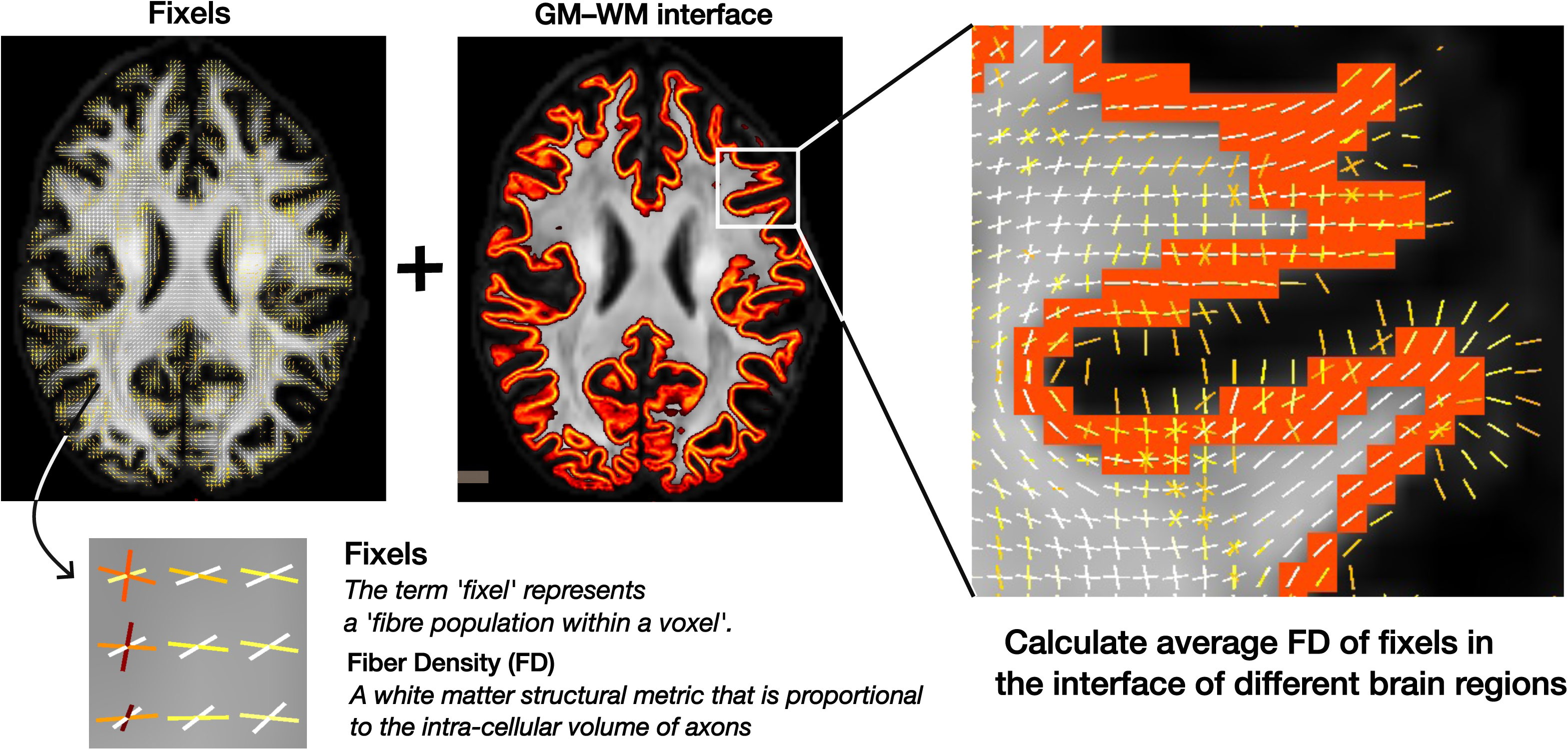
Overview of the analysis of white matter structure at the grey matter–white matter (GM–WM). This figure illustrates the process of analyzing white matter at the GM–WM interface. Fixels represent individual fiber populations within a voxel. Based on these fixel-level metrics, the average fiber density (FD) at the interface adjacent to each region of interest (ROI) was calculated.

FD was calculated in MRtrix3 (version 3.0.3) following the initial steps of the standard FBA pipeline. Response functions for GM, WM, and CSF were estimated from the preprocessed diffusion data using Dhollander’s unsupervised response function estimation algorithm (dwi2response) (45,46). Fiber orientation distributions were computed using MSMT-CSD (dwi2fod) based on group-average response functions, followed by bias-field correction and global intensity normalization (mtnormalise). Finally, FD was quantified via fixel segmentation (fod2fixel) (31).

The GM–WM interface was defined using the *5tt2gmwmi* command in MRtrix3 (version 3.0.3) (47). This command identified the transition zone based on the partial volume fractions of GM and WM in each voxel, which were computed from T1-weighted images using the FMRIB Automated Segmentation Tool (47,48).

For the main analysis, region of interest (ROI)-level analyses in the GM–WM interface were conducted in each subject’s native space. The ROI-based analysis followed a pipeline similar to our previous study (16). ROIs were defined based on the Destrieux atlas, and the adjacent GM–WM interface was assigned to each ROI. We calculated the average FD of the fixels within each region. For comparison with FD, fractional anisotropy (FA) was estimated using the *dtifit* tool in FSL (v6.0.4), excluding the b = 3000 s/mm² shell to ensure appropriate tensor model fitting (49).

The effect of the scanner and protocol differences on the variables of cortex and WM was reduced by ComBat harmonization (50–52) (Figure S2). ComBat was applied to the mean ROI values of each measure, with age, sex, and diagnosis specified as biological covariates to be preserved.

### Spatial correlation of group difference effect size between FD of the GM–WM interface and cortical thickness and surface area

We assessed the spatial correspondence between effect size maps of FD and cortical features using a spin-test implemented in the ENIGMA toolbox (53,54). For each feature, regional effect sizes (Cohen’s *d*) from the Destrieux atlas were projected onto the Freesurfer average surface, which contains 10,242 vertices per hemisphere, by nearest-neighbor interpolation. The observed spatial correlation was compared against a null distribution generated from 10,000 spherical rotations, and spin-based permutation p-values were computed. In each rotation, one hemisphere was randomly rotated, and the other hemisphere was rotated using the mirrored transformation to preserve interhemispheric symmetry. As supplementary analyses, we examined spatial correlations between cortical thickness and surface area, between FD and FA, and between FA and cortical measures.

### Post hoc analysis 1: Template-space analysis of fixel-based metrics using a multimodal FOD template

To reduce the influence of inter-individual variability in ROI definition and enable comparisons within spatially matched ROIs, we performed complementary analyses in template space using a multimodal FOD template (55). This multimodal template provides higher spatial specificity and sharper delineation of the GM–WM interface. FOD images were non-linearly registered to the template, and the GM–WM interface was defined and assigned to Destrieux atlas regions for group comparisons. In addition to FD, as supplementary analyses, fiber cross-section (FC) and fiber density and cross-section (FDC) were also quantified (43). Detailed procedures are provided in the Supplementary Methods.

### Post hoc analysis 2: Layer-specific characterization of FD across the GM–WM interface

To further characterize FD across the GM–WM interface, we performed additional layer-specific analyses (Figure S3). The GM–WM interface was subdivided into a GM-side layer and a WM-side layer based on FAST-derived partial volume fractions. In addition, an extra WM layer located one voxel beneath the interface was examined.

### Statistical analysis

Demographic, clinical, and cognitive indices were compared between groups using the two-sample t-test, Mann-Whitney U test, and chi-square test. FD and FA were compared between groups using linear regression, with sex, age, and head motion as covariates. Cortical thickness and surface area were adjusted for sex and age. Brain metrics were assessed across multiple brain regions with the False Discovery Rate (FDR) correction (56). Group-by-sex and group-by-age interaction terms were also tested.

We also assessed the influence of GM proportion within each ROI. The differences in GM proportion could potentially influence FD values, as a higher proportion of GM within a voxel may reduce FD due to partial volume effects, even though MSMT-CSD separates WM-like and GM-like diffusion signals. We performed independent-samples t-tests to assess group differences in GM proportion across ROIs. We also examined the association between group differences in GM proportion and FD using Pearson correlations of effect sizes. GM proportion was included as an additional covariate in the linear model to assess group differences in FD.

In post hoc analysis 1, intracranial volume was additionally included as a covariate for FC and FDC. In post hoc analysis 2, regional GM proportion was additionally included as a covariate to account for partial-volume effects for the GM- and WM-side layers of the interface.

All analyses were conducted using R statistical software (version 4.2.3), using packages including the *car* package (57,58).

### Association between FD and clinical symptom scores and antipsychotic dosage

An exploratory analysis was conducted to examine the relationship between PANSS subscales and average FD at the GM–WM interface across the whole brain in individuals with early psychosis, controlling for age and sex (37). The same models tested the associations with antipsychotic medication dosage, calculated as chlorpromazine equivalents (mg/day).

## RESULTS

### Clinical and demographic characteristics of the participants

Early psychosis and control groups were matched in sex (both 70.5% male), age, and race (Table 1).

**Table 1.**
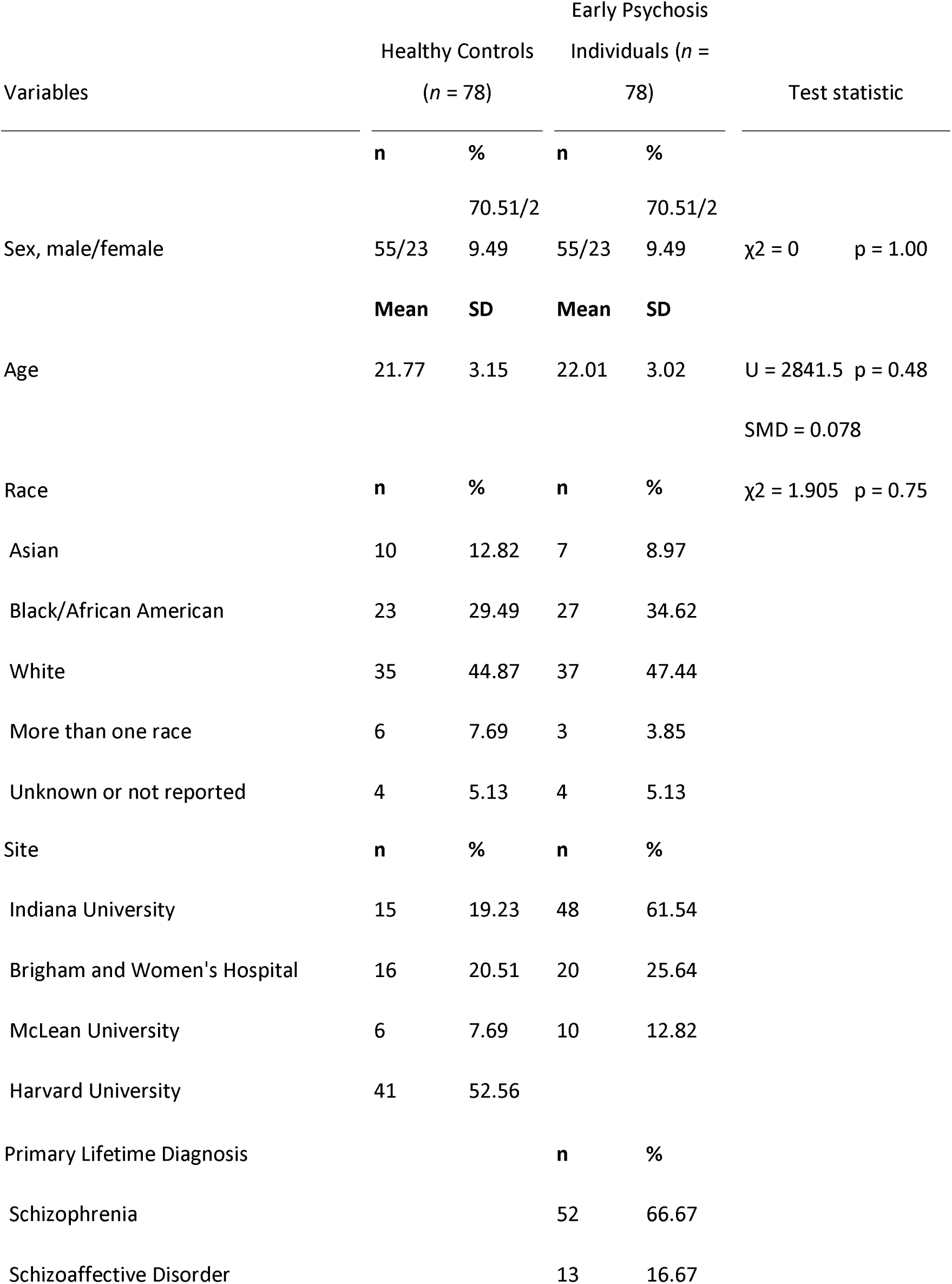
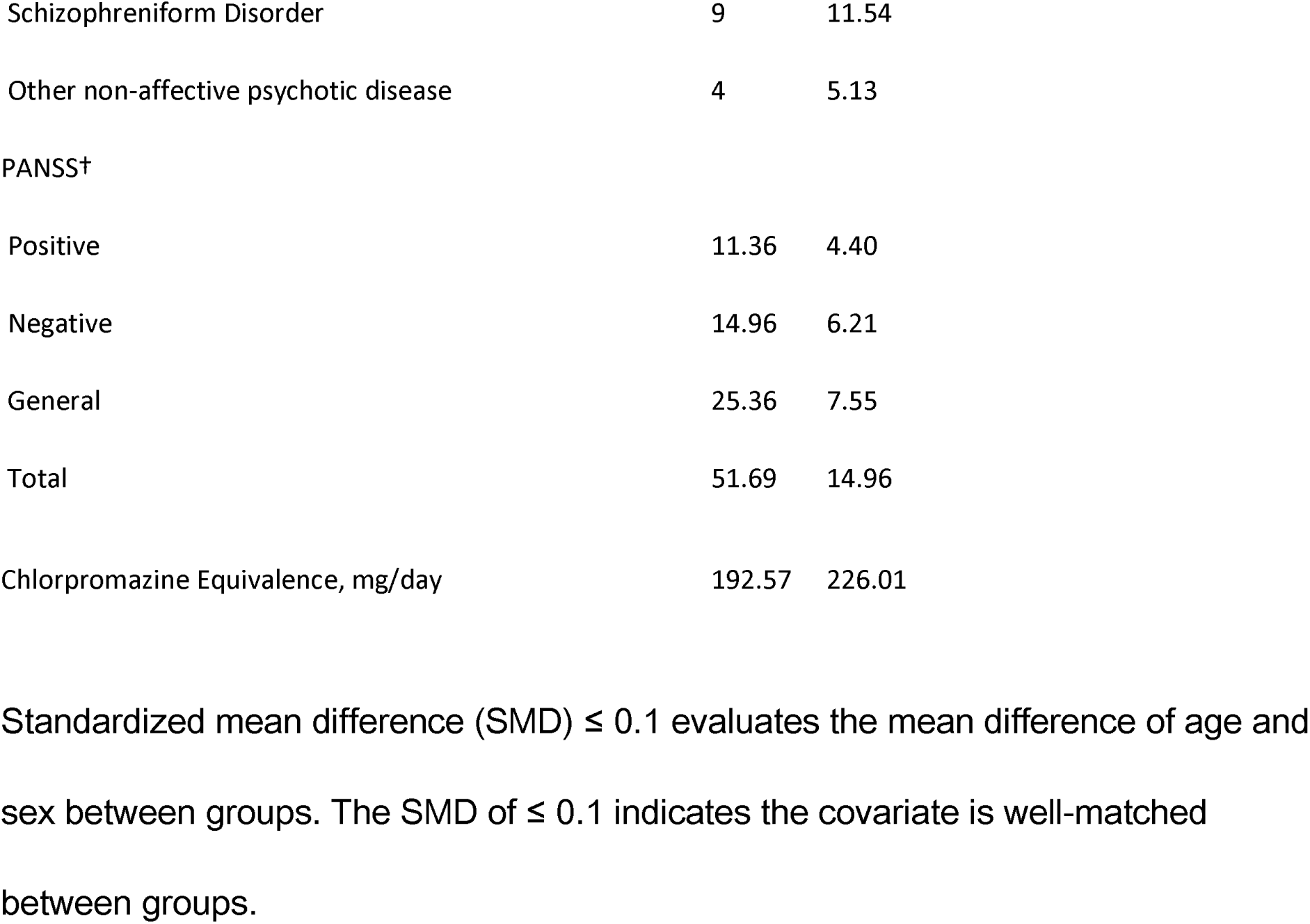
Demographic and clinical data.

### Group differences in cortical GM structural metrics

In early psychosis, widespread reductions in cortical thickness were observed relative to healthy controls, predominantly in temporal, frontal, and insular regions. No significant differences were observed in cortical surface area.

### Group differences in WM microstructure of the GM–WM interface

Contrary to expectation, no significant differences in FD were observed across ROIs. Regions that were significant at an uncorrected threshold are provided in the Supplementary Materials (Figure S4). No significant group-by-sex or group-by-age interactions were observed. FA, included as a complementary metric, also showed no significant group differences.

Some ROIs exhibited lower GM proportion in the healthy control group, and greater between-group differences in GM proportion were associated with smaller between-group differences in FD (Figure S5). We repeated the analysis including GM proportion as an additional covariate in the model; no significant group differences in FD were observed.

### Spatial correlation of group difference effect sizes between FD of the GM–WM interface and cortical thickness and surface area

We examined the spatial correlations between the effect sizes of group differences in FD and those in cortical measures across brain regions using a spin test (Figure 4). There was a significant positive correlation between effect sizes of group differences in FD and cortical thickness (*r* = 0.26, *p* = 0.011) and between FD and cortical surface area (*r* = 0.28, *p* = 0.012).

**Figure 3.**
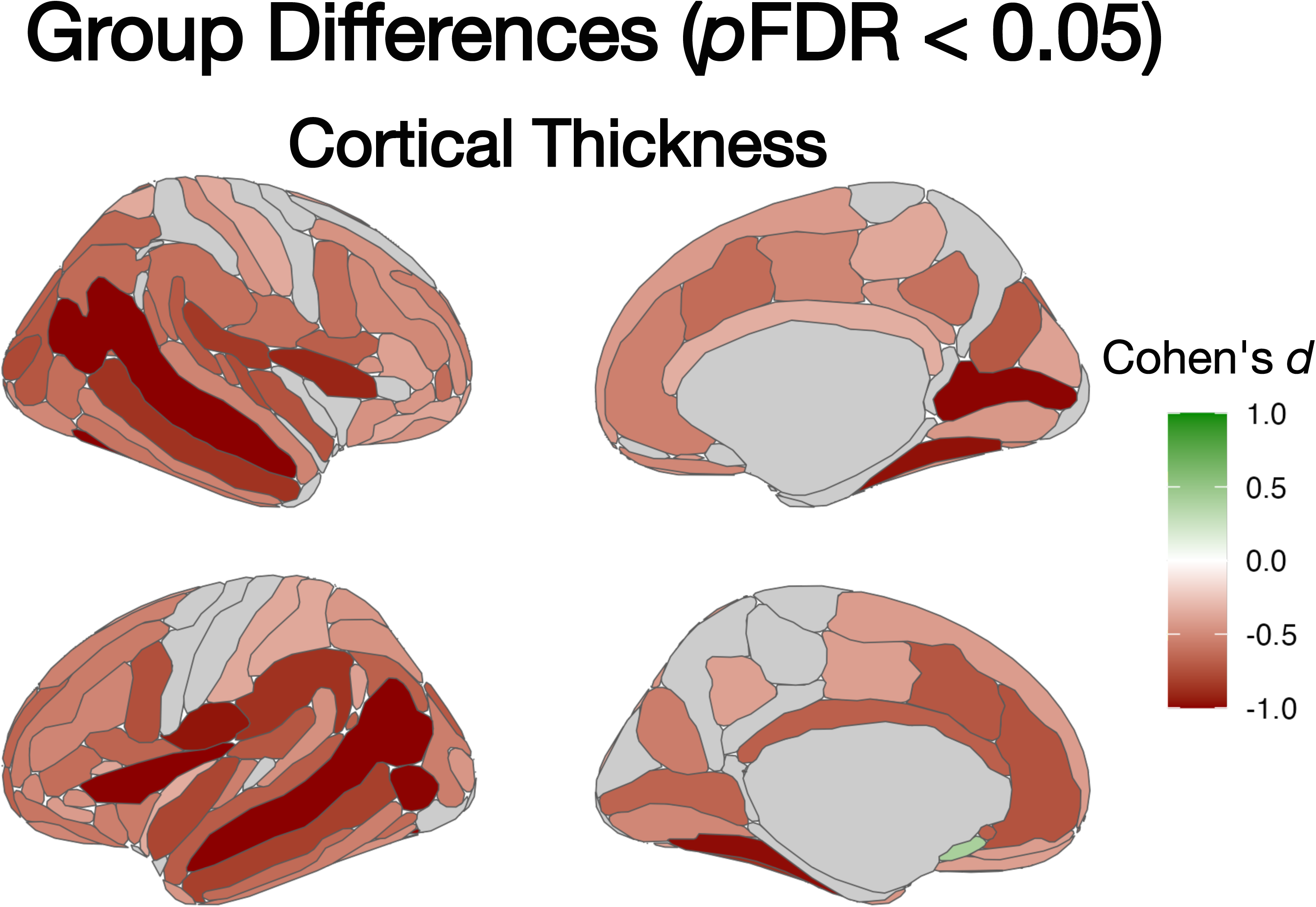
Group differences in cortical thickness. Group differences in cortical thickness were assessed using regression models with age and sex as covariates. Effect sizes for significant differences in cortical thickness between individuals with early psychosis and healthy controls are shown (*p*FDR < 0.05). Red indicates regions where cortical thickness is lower in individuals with early psychosis, whereas green indicates regions where cortical thickness is lower in healthy controls.

**Figure 4.**
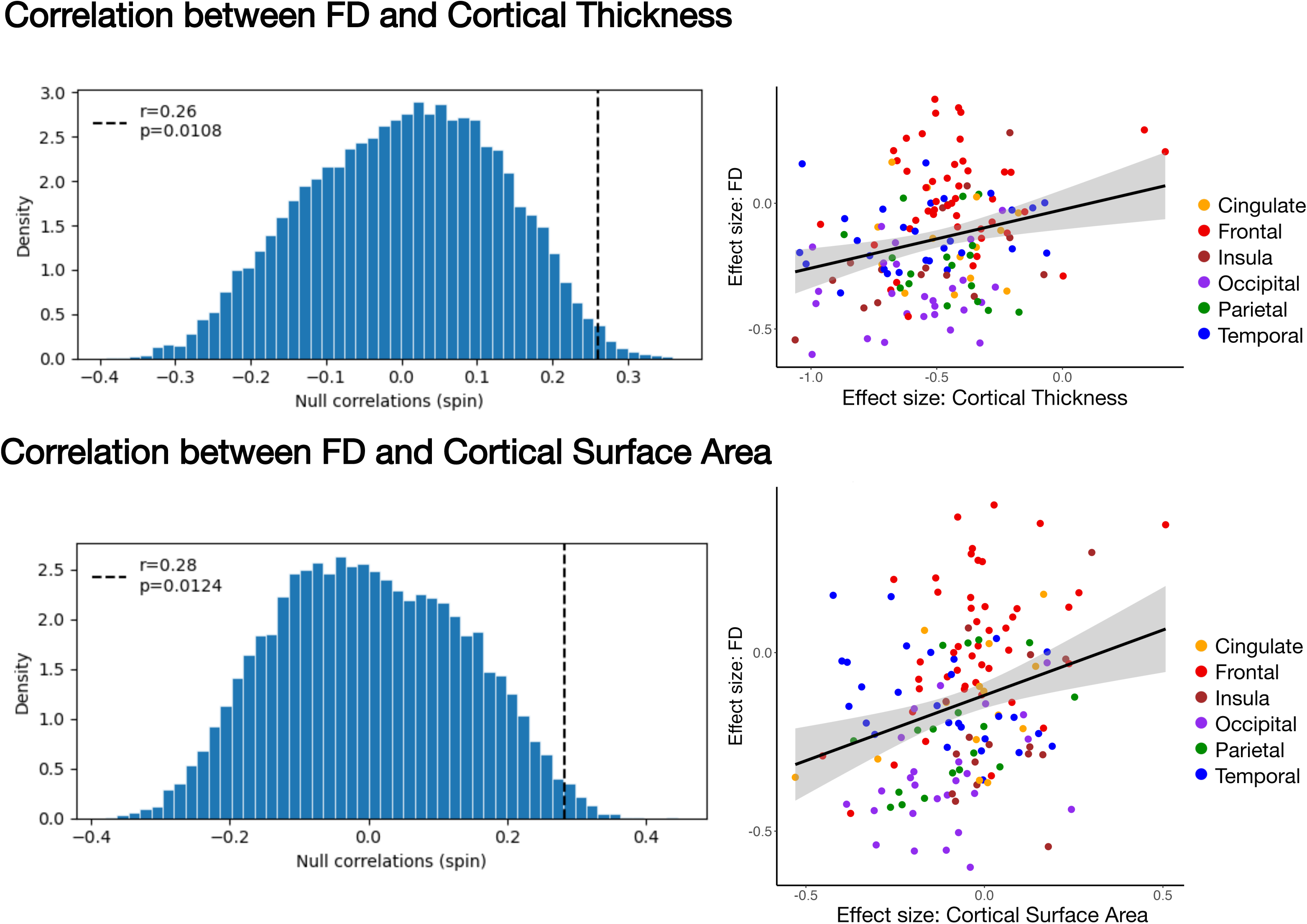
Spatial correlations between fiber density (FD) and cortical thickness and surface area. Spatial correlations between regional effect sizes of fiber density and cortical thickness (top) and cortical surface area (bottom) are shown. The left panels display the null distributions of spatial correlations generated by a spin test, with the vertical dashed line indicating the observed correlation. The right panels show scatter plots of the observed regional effect sizes with a fitted regression line. Each point represents a brain region, and colors indicate anatomical regions.

Additional analyses showed no significant correlation between effect sizes in cortical thickness and surface area (*r* = -0.04, *p* = 0.34), whereas those in FD and FA were strongly correlated (*r* = 0.76, *p* < 0.0001). FA showed significant spatial correlations with both cortical thickness (r = 0.22, p = 0.019) and surface area (r = 0.30, p = 0.0037) (Figure S6).

### Post hoc analysis 1: Template-space analysis of fixel-based metrics using a multimodal FOD template

Analyses performed in template space using a common ROI definition across all participants showed no significant group differences in FD in any of the GM–WM interface ROIs. Supplementary analyses of fiber cross-section (FC) and fiber density and cross-section (FDC) also showed no significant group differences.

As averaging FD across all fixels within each ROI may reduce sensitivity to specific fiber populations, we conducted an exploratory fixel-level analysis of FD in regions showing uncorrected significance in the template-space analysis, using MRtrix3 *fixelcfestats* with age, sex, and in-scanner head motion as covariates (42). We focused on the left superior and inferior segments of the circular sulcus of the insula, which showed the largest effect sizes (superior: *d* = −0.42, uncorrected *p* = 0.0093, *p*FDR = 0.52; inferior: *d* = −0.47, uncorrected *p* = 0.021, *p*FDR = 0.52). This analysis identified significantly reduced FD in superficial white matter tracts extending from the inferior frontal gyrus to the insular region (Figure S7).

### Post hoc analysis 2: Layer-specific characterization of FD across the GM–WM interface

On the GM-side of the interface, a significant reduction in FD was observed in a limited occipital region (left anterior occipital sulcus and preoccipital notch: *d* = −0.61, *p*FDR = 0.045), while significant increases in FD were identified in frontal regions (left lateral orbital sulcus: *d* = 0.53, *p*FDR = 0.045; right frontomarginal gyrus and sulcus: *d* = 0.55, *p*FDR = 0.045) (Figure S8). On the WM-side of the interface, a significant reduction in FD was detected in the right middle occipital sulcus and lunatus sulcus (*d* = −0.69, *p*FDR = 0.021). No regions showed significant group differences in the sub-interface layer.

### Association between FD at the GM–WM interface and symptom severity and antipsychotic dosage

No significant associations were found between FD at the interface and symptom severity or antipsychotic dosage.

## DISCUSSION

In this study, we aimed to investigate structural differences in the grey matter–white matter (GM–WM) interface and associations with cortical morphology in early psychosis. There were two primary findings: (1) no significant alterations were observed in fiber density (FD) despite widespread cortical thinning; and (2) the spatial distribution of FD effect sizes showed significant positive correlations with cortical thickness and surface area. By conducting complementary post hoc analyses, we also demonstrated a marked contrast between prominent cortical GM alterations and the lack of detectable group differences in WM at the GM–WM interface.

Previous studies using the same early psychosis cohort reported subtle or no WM alterations in deep WM tracts (59,60). Even when the interface was separated into GM-and WM-side layers and deeper layers were examined, group differences were limited or absent. The marked contrast between widespread cortical thinning and the absence of WM differences at the interface in early psychosis is consistent with findings across stages of illness proposing that GM reduction emerges earlier than WM alterations (15). The central involvement of GM pathology in the early stages of schizophrenia-spectrum disorders is supported by evidence that acceleration of cortical thinning prior to onset, rather than white matter alterations, predicts transition to psychosis (61–63). As WM changes are believed to progress over time in schizophrenia-spectrum disorders, changes at the GM–WM interface may become more detectable in later stages of illness, rather than at this ‘early psychosis’ stage. Post-mortem studies have consistently identified cortical alterations, such as dendritic spine loss, reduced synaptic density, and decreased neuronal soma size, as primary changes, which may subsequently influence axonal integrity at the GM–WM interface (64–71).

Alternatively, given the limited evidence for neurodegeneration and the accumulating evidence of glial alterations, including oligodendrocyte abnormalities, FD, which is sensitive to intra-axonal water diffusion, may not necessarily show disease-specific alterations in schizophrenia-spectrum disorders (31,44,72,73). Post-mortem and MRI studies have reported myelin-related changes in schizophrenia, and WM abnormalities identified in previous studies using tensor-derived metrics in schizophrenia-spectrum disorders may not reflect axonal degeneration, but rather other WM alterations, such as myelin-related changes (18,74–77).

Although no group differences were observed at the GM–WM interface, the significant positive association between effect size maps of cortical GM and WM at the interface suggests a relationship between the two. While a previous study reported a negative association, differences in findings may be attributable to differences in illness stage, depth from the cortex, or methodological approaches (16). The finding that FD spatial patterns were associated with both cortical thickness and surface area is interesting, as these cortical metrics reflect different aspects of cortical organization at structural, genetic, and developmental levels (78–84). These results suggest that multiple aspects of cortical alterations are associated with the adjacent WM structure. Although no significant group differences were detected, these findings raise the possibility that subtle WM changes may already be emerging at this early stage of illness.

Methodologically, the implementation of MSMT-CSD allowed us to study white matter microstructure at the GM–WM interface by separating different tissue compartments from the diffusion signal (30). However, the fiber density (FD) measure still depends upon the level of white matter signal within a given voxel, which can be impacted by the presence of other tissue types (in this case, the presence of GM). As such, we included additional analyses incorporating the GM proportion within each ROI as a covariate, along with analyses performed in a robust template space, to minimize the impact of individual differences in GM proportion within defined ROIs. The latter analysis used a multimodal template that could more precisely delineate the GM–WM interface than one generated using DWI-derived FODs only (55). Despite these different approaches, we consistently found no evidence for differences in mean FD in GM–WM interface ROIs. Taken together, these analyses suggest that WM structure, even at fiber entry points adjacent to the cortex, is relatively preserved during the early stages of psychosis.

This study has some limitations. The cross-sectional design precludes causal inferences about the relationships between GM and WM. Illness duration was unavailable in the dataset, limiting assessment of illness course. While no associations with antipsychotic dosage were found, studies in antipsychotic-naive individuals are needed to rule out potential confounding effects of medication. In addition, because the GM–WM interface includes both superficial white matter fibers and long-range projection fibers, ROI-level averaging of FD across crossing-fiber fixels could not distinguish between these fiber populations (26). This averaging did not capture fixel-level WM structure and may contribute to the strong association between FD and FA (Figure S6). Indeed, our exploratory fixel-level analysis suggested that alterations may occur within specific fiber populations in superficial white matter (Figure S7). Future work that specifically examines superficial U-fibers may be insightful. Finally, considering the heterogeneity in schizophrenia-spectrum disorders, especially in the early stages (85–88), larger samples are needed to validate our findings.

## Conclusion

In early psychosis, widespread cortical thinning was observed, whereas WM microstructure at the GM–WM interface was relatively preserved. However, the spatial pattern of group differences in FD at the interface corresponded to cortical morphological changes, suggesting that cortical alterations in early psychosis are linked to the underlying WM structure.

## Data Availability

Brain imaging and cognitive and clinical data from the Human Connectome Project for early psychosis can be requested from https://www.humanconnectome.org/study/human-connectome-project-for-early-psychosis. Brain imaging and cognitive data from the Human Connectome Project Development can be requested from https://www.humanconnectome.org/study/hcp-lifespan-development. Access to these datasets requires approval. Other data can be obtained from the corresponding author upon reasonable request.

https://www.humanconnectome.org/study/hcp-lifespan-development

## ACKNOWLEDGEMENTS

Research using Human Connectome Project for Early Psychosis (HCP-EP) data reported in this publication was supported by the National Institute of Mental Health of the National Institutes of Health under Award Number U01MH109977. The HCP-EP 1.1 Release data used in this report came from DOI: 10.15154/1522899.

Y.S. was supported by the Research Training Program Scholarship, and Japan Student Services Organization Scholarship. M.R.-C. was supported by a Ramon y Cajal Research Fellowship (RYC-2017-23144), Spanish Ministry of Science, Innovation and Universities and an AEI Consolidator Grant (CNS2023-144038) from the State Research Agency Spanish Ministry of Science, Innovation and Universities; she was partially supported by the Instituto de Salud Carlos III, ISCIII, (PI18/00753, PI21/00701, PI24/01298), CIBER -Consorcio Centro de Investigación Biomédica en Red-(CB/07/09/0023), Spanish Ministry of Science and Innova Spanish Ministry of Science, Innovation and Universities, cofinanced by ERDF Funds from the European Commission, “A way of making Europe”, by the European Union, Madrid Regional Government (S2022/BMD-7216 AGES 3-CM), European Union Structural Funds, European Union Seventh Framework Programme and European Union H2020 Programme, Fundación Familia Alonso and Fundación Alicia Koplowitz. M.A.D. was supported by Al & Val Rosenstrauss Fellowship 2025 from the Rebecca L Cooper Foundation. R.M. is the recipient of an Australian Research Council Discovery Early Career Researcher Award (ID: DE240101035). C.P. was supported by a National Health and Medical Research Council (NHMRC) Investigator Grant (ID: 1196508) and NHMRC Program Grant (ID: 1150083). C.W. was supported by an NHMRC) Investigator Grant (ID: 2034232).

This manuscript was posted as a preprint on medRxiv (DOI: https://doi.org/10.1101/2025.05.13.25327498).

## Code Availability

The code developed and used for the analyses in this study has been made openly available on GitHub at https://github.com/YSaito00/GMWMinterface_CSD_analysis.

## Disclosures

The authors declare no competing interests.

## Notes

### Competing Interest Statement

The authors have declared no competing interest.

### Author Declarations

The study used (or will use) ONLY openly available brain imaging and cognitive and clinical data from the Human Connectome Project for early psychosis (https://www.humanconnectome.org/study/human-connectome-project-for-early-psychosis) and brain imaging and cognitive data from the Human Connectome Project Development (https://www.humanconnectome.org/study/hcp-lifespan-development).

### Summary of Updates

This manuscript represents a revised version of a previously submitted study, originally entitled "Interrogating the grey-white matter boundary in early psychosis: fiber-specific reductions in superficial white matter." Following peer review, we identified an inconsistency in the estimation of response functions used for fiber density (FD) calculation. To ensure methodological consistency across participants, we reprocessed the diffusion MRI data, which led to changes in the primary findings. In addition, we conducted complementary post hoc analyses to more rigorously evaluate the results.

