## Supplementary Material for "Preserved white matter structure at the grey matter–white matter interface despite widespread cortical thinning in early psychosis"

### **Supplemental Information**

### **MATERIALS AND METHODS**

#### ***Participants***

The study consisted of early psychosis individuals from the Human Connectome Project for Early Psychosis (HCP-EP) and healthy controls from two datasets, HCP-EP and the Human Connectome Project in Development (HCP-D) (1,2). Due to differences in the inclusion and exclusion criteria for healthy controls between the HCP datasets, we included those from HCP-D who did not take psychiatric medication and did not have parents diagnosed with schizophrenia spectrum disorders. Considering the typical period of psychosis onset, we included subjects between the ages of 16 and 30 (3). We included subjects only from Harvard University out of the four imaging sites of HCP-D to minimize non-biological variability caused by imaging site differences. As a result, subjects between the ages of 16 and 30 with T1-weighted and diffusion-weighted images with no quality issues in the image processing stage were selected: 78 early psychosis individuals and 45 healthy controls from HCP-EP, and 74 healthy controls from HCP-D (Figure S1).

To match the number of subjects, sex, and age between groups, propensity score matching was performed on pooled controls (4,5). We calculated the propensity score of each subject using a logistic regression model with sex and age as covariates and performed a nearest-neighbor matching algorithm without replacement based on the propensity score. We confirmed the matching achieved adequate balance by verifying that the standardized mean differences of sex and age were below 0.1 (Table 1).

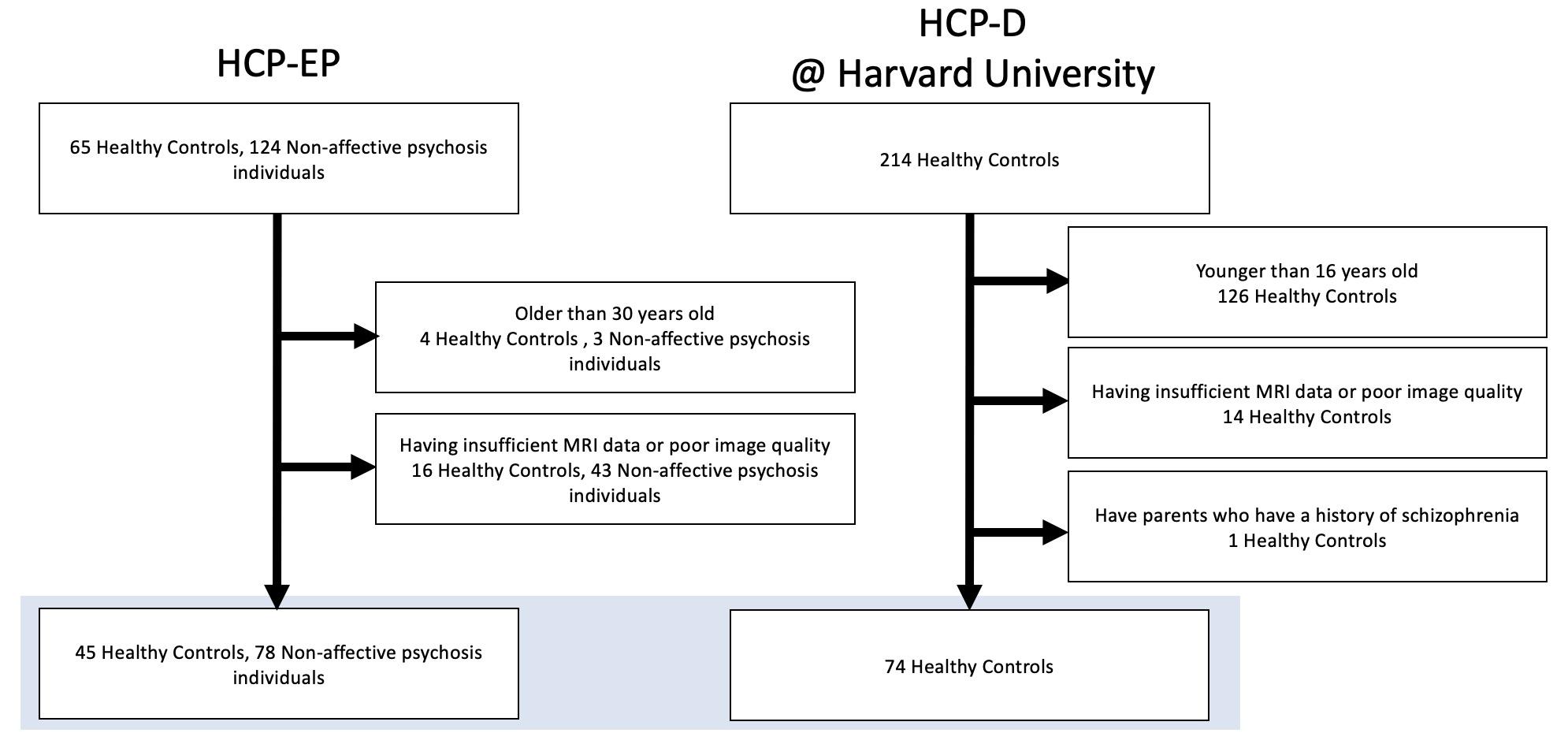

Figure S1. Flowchart of subject selection. The study selected subjects who provided sufficient MRI data and were aged between 16 and 30 years.

#### Harmonization

We harmonized all the variables across four different imaging sites to reduce the effect of the scanner and protocol differences. Variables were harmonized using ComBat, which maintains biological variations related to diagnosis, sex, and age in diffusion MRI measures (6–8). ComBat reduced significant differences in all brain regions across these imaging sites (Figure S2) (p < 0.05).

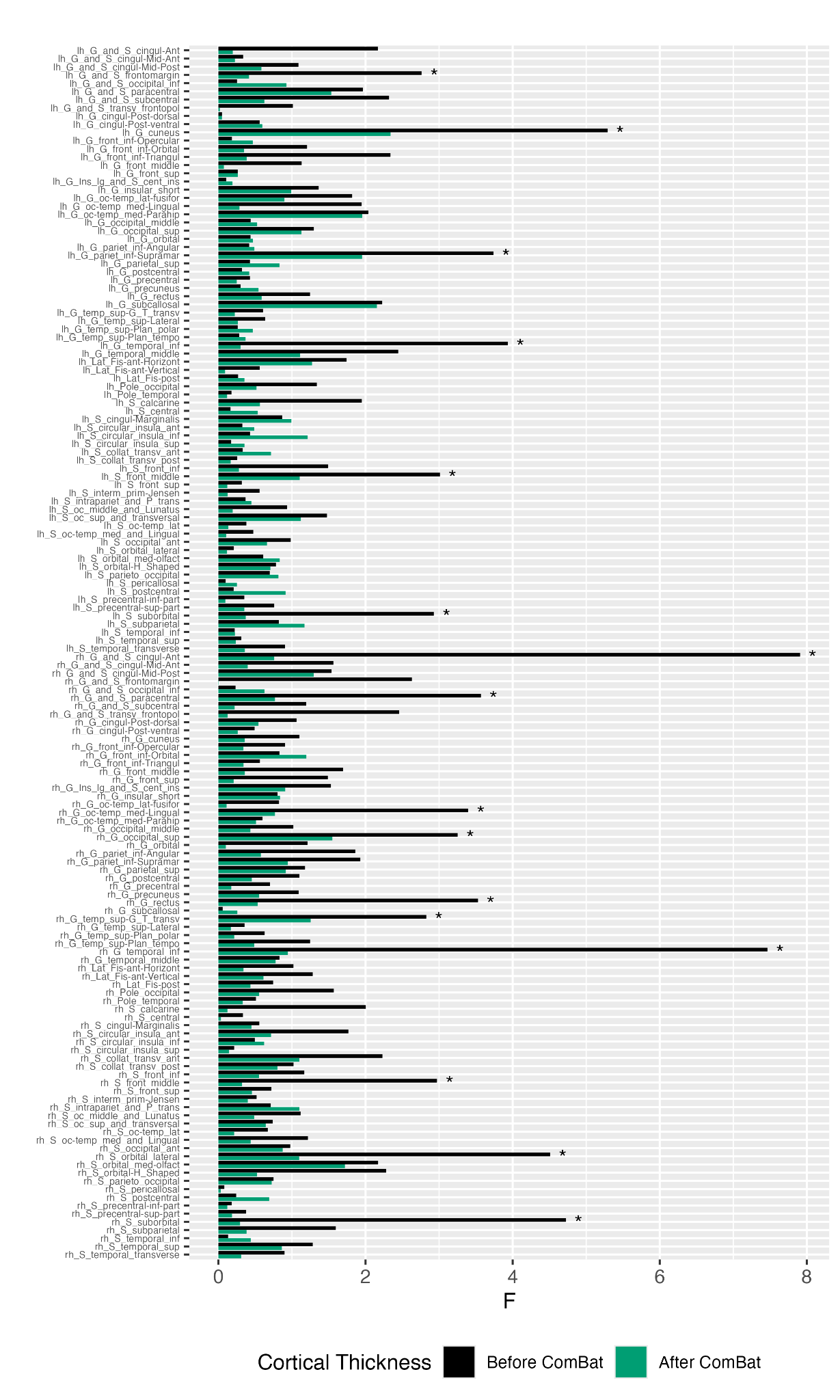

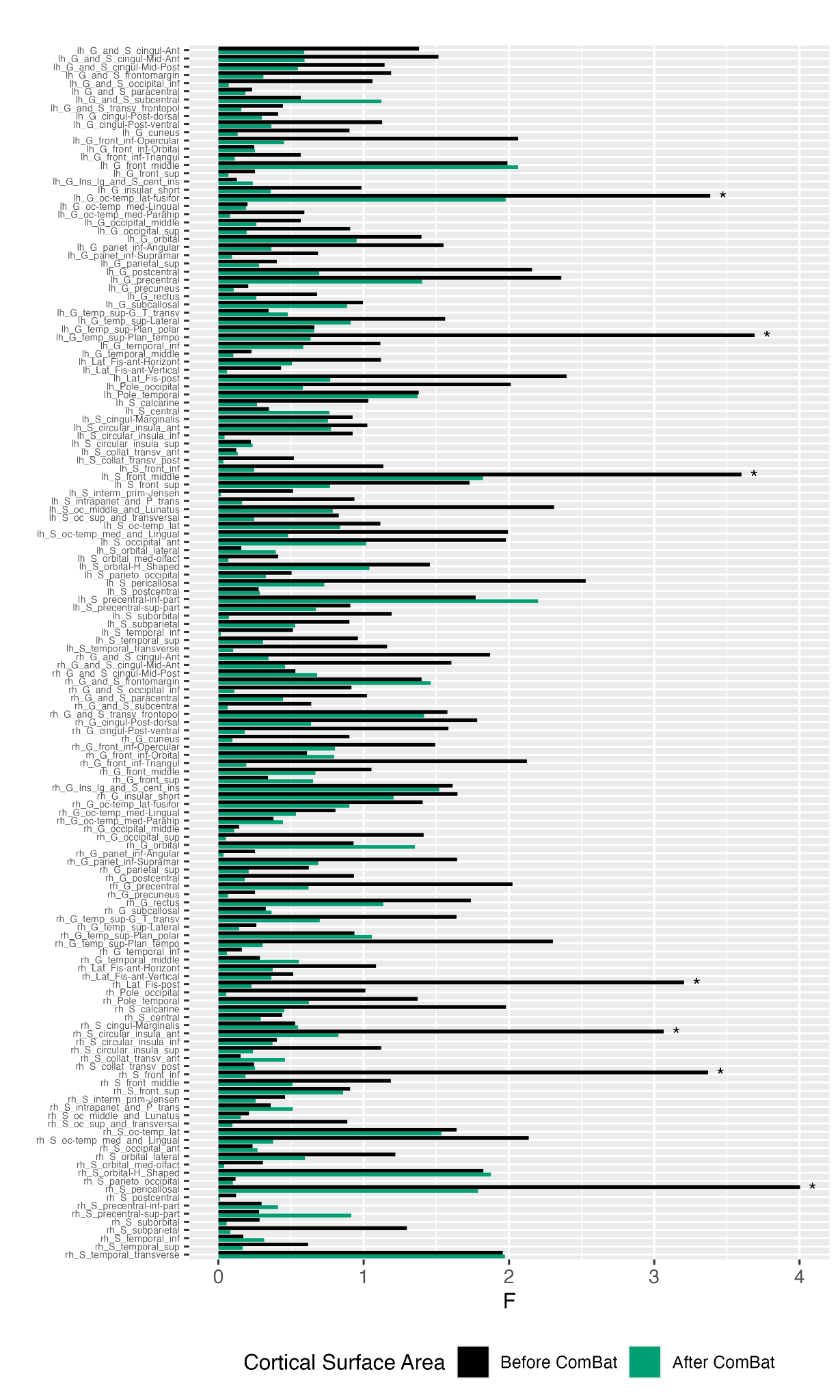

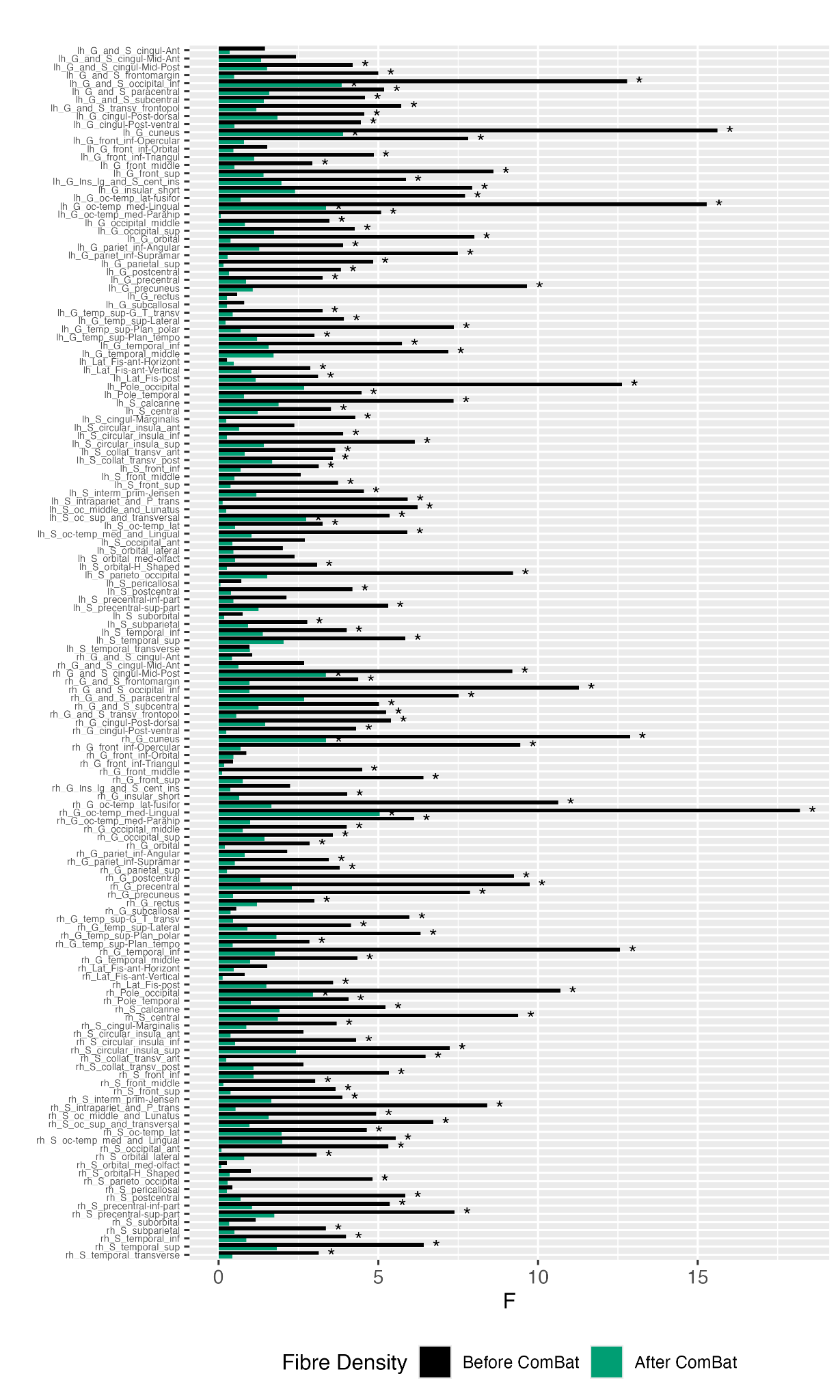

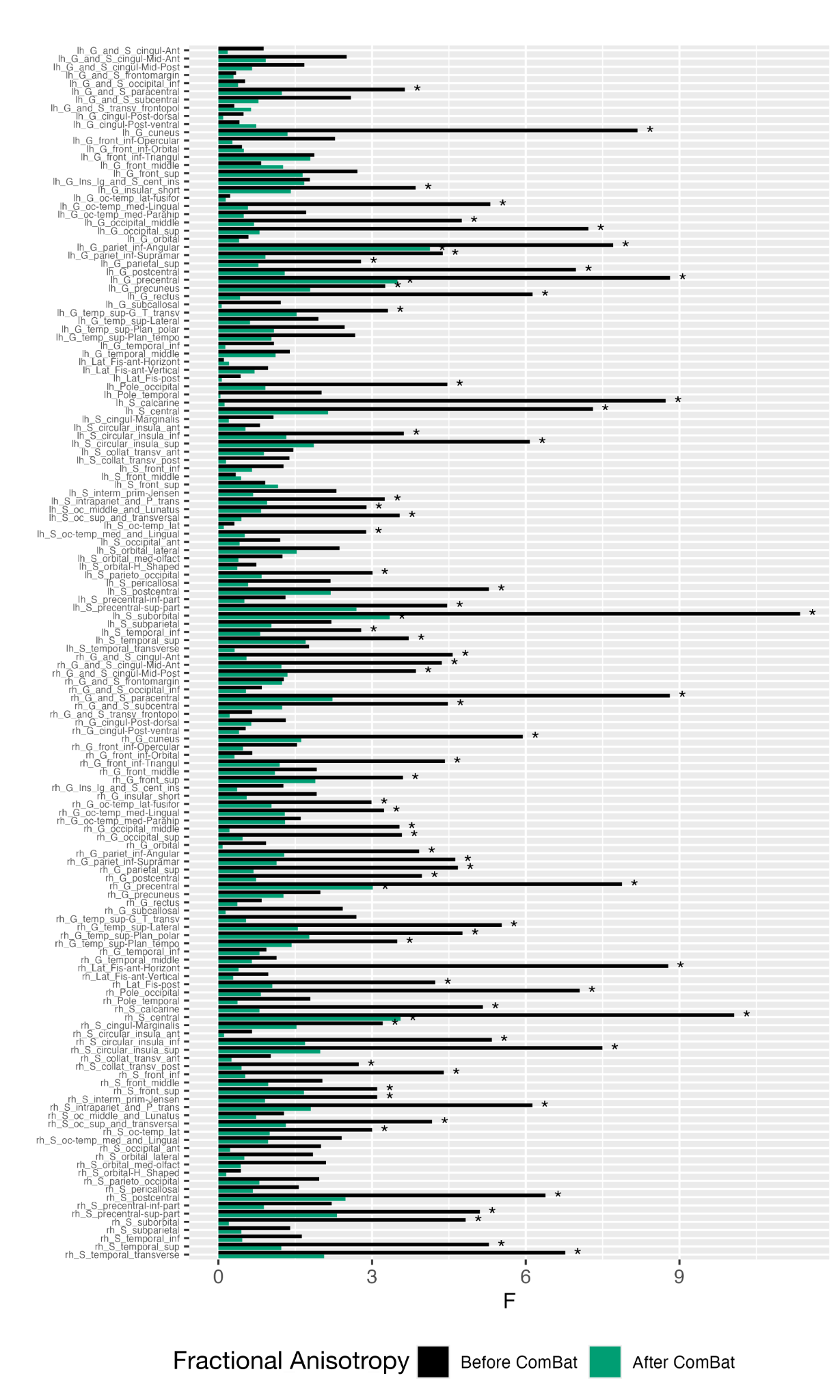

Figure S2. Harmonization of cortical thickness, surface area, fiber density, and fractional anisotropy. *F*-values decreased across the variables after ComBat harmonization. Asterisks display significant differences between scanners (*p* < 0.05).

***Template-space analysis of fixel-based metrics using a multimodal FOD template***

To evaluate whether the observed findings were driven by inter-individual variability in ROI definition in subject space and to enable group comparisons within spatially matched ROIs across subjects, we performed complementary analyses in template space using a multimodal FOD template (9). This multimodal template provides improved spatial specificity and sharper delineation of the GM–WM interface. A multimodal brain template was constructed using 25 age- and sex-matched healthy controls (age 22.0 ± 4.09 [SD], 60% male) and 25 individuals with early psychosis (age 22.6 ± 3.04 [SD], 64% male). Template generation was performed using multivariate symmetric group-wise normalization applied to T1-weighted and T2-weighted images, and diffusion MRI–derived FA and mean diffusivity (MD), implemented in Advanced Normalization Tools (ANTs, version 2.4.3) (10). After initial linear alignment, all images were iteratively warped into a common space using symmetric group-wise normalization, optimizing the template to minimize differences between the template and individual subjects. Template updating, including appearance and shape optimization, was performed as an iterative procedure and set to converge within a maximum of four iterations.

The preprocessed diffusion-weighted images were upsampled to 1.25 mm isotropic resolution to improve delineation of the GM–WM interface. As in the subject-space analyses, FOD images were estimated using MSMT-CSD with a common set of group-averaged response functions. We applied the estimated ANTs transformations to each individual’s FOD image and generated the FOD template by voxel-wise averaging of the warped FOD images across subjects. After non-linear registration of all participants’ FOD images to the template space, the GM–WM interface was defined using the 5tt2gmwmi command based on the template derived from T1-weighted images, following the same procedure as in subject space. The GM–WM interface was assigned to ROIs defined by the Destrieux atlas, enabling group comparisons based on shared ROIs across participants.

As supplementary analyses in template space, fiber cross-section (FC) and the combined measure of fiber density and cross-section (FDC) were also quantified following the standard FBA pipeline (11). FC captures macroscopic differences in fiber-bundle morphology, whereas FDC combines microscopic fiber density and macroscopic bundle morphology to provide a more comprehensive index of intra-axonal content.

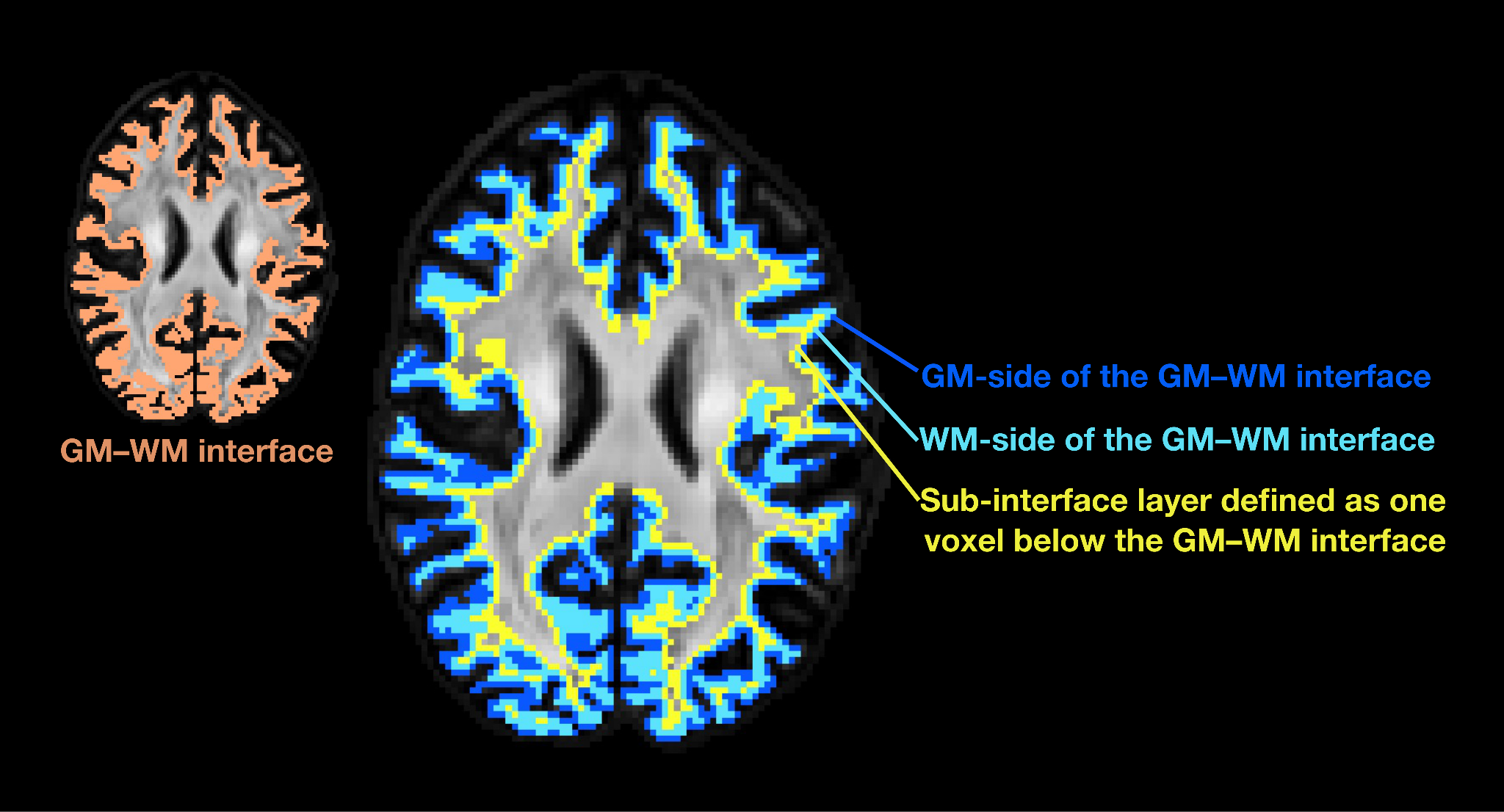

Figure S3. The GM–WM interface was subdivided into a GM-side layer and a WM-side layer based on FAST-derived partial volume fractions. Voxels with a higher GM fraction were classified as the GM-side layer (blue), while voxels with a higher WM fraction were classified as the WM-side layer (light blue). In addition, a deeper WM layer was defined as the region located one voxel beneath the GM–WM interface within the WM (yellow). This layer was defined by shifting the interface one voxel (1.5 mm) toward the WM, thereby including voxels directly adjacent to the interface. As a result, the layer covered WM located approximately 1.5 to 2.6 mm below the GM–WM interface.

**RESULTS**

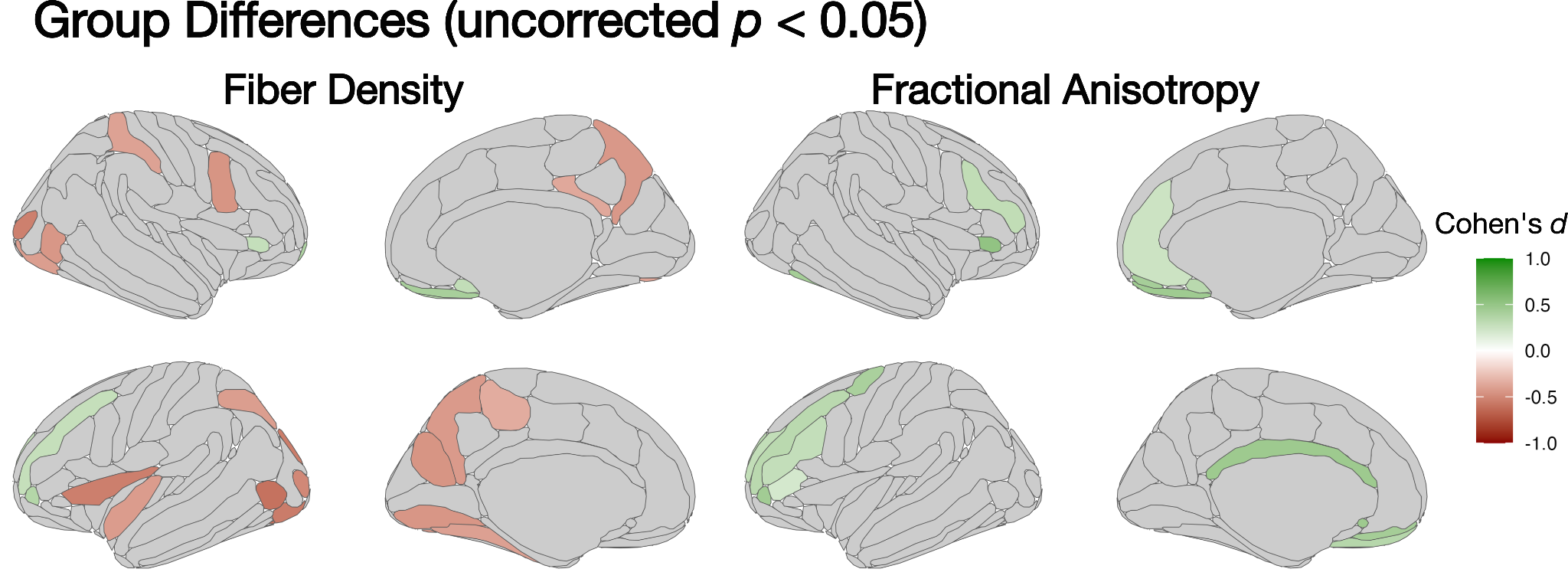

Figure S4. Group differences in FD of the GM–WM interface. Group differences in FD were assessed using regression models with age, sex, and head motion as covariates. Brain regions showed significant differences in FD between individuals with early psychosis and healthy controls (*p* < 0.05). Red indicates regions where FD is lower in individuals with early psychosis, while green indicates regions where FD is lower in healthy controls.

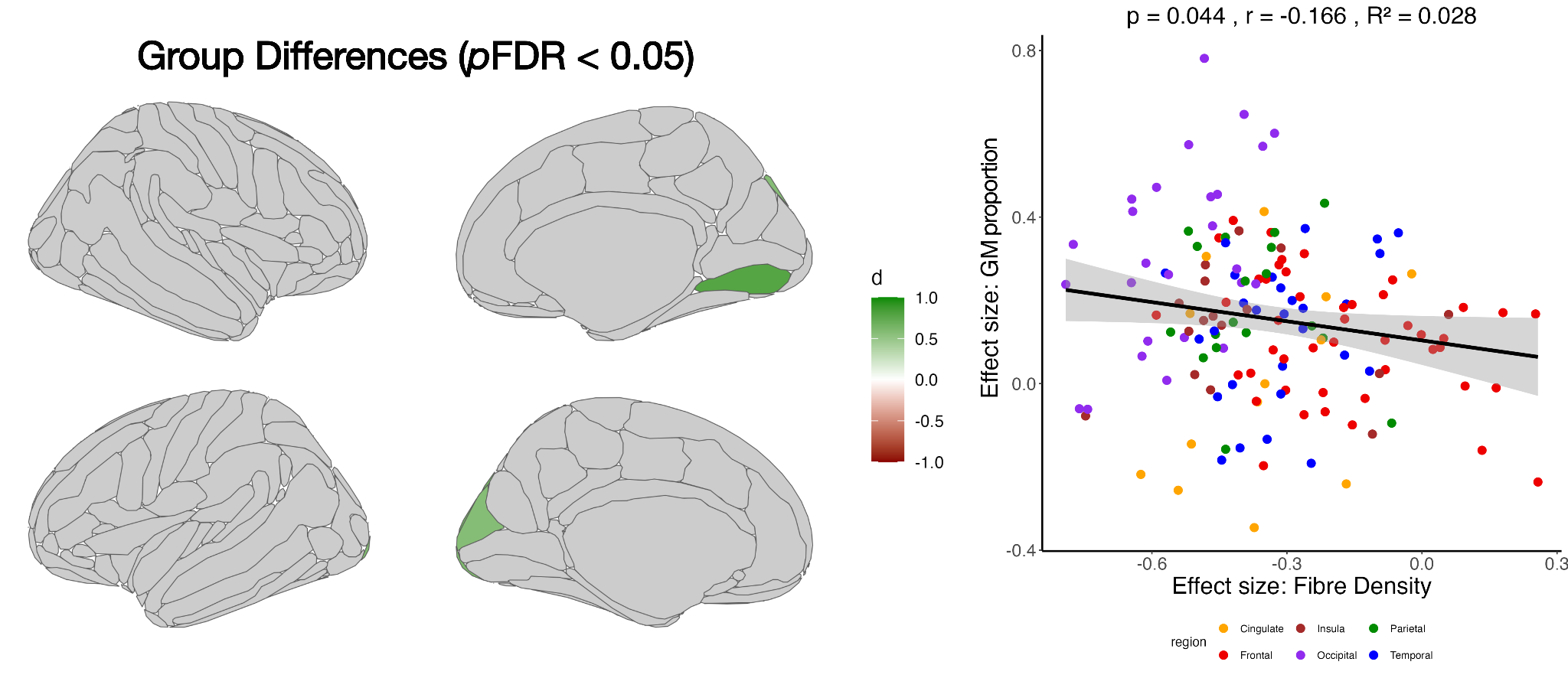

Figure S5. Group differences in GM proportion of the GM–WM interface in subject space.

The left panel displays group differences in GM proportion assessed using independent-samples t-tests. Brain regions showing significant differences between individuals with early psychosis and healthy controls (pFDR < 0.05) are highlighted. The right panel shows a scatter plot of the observed regional effect sizes with a fitted regression line, explaining the relationship between effect sizes of GM proportion and those of FD. Each point represents a brain region, and colors denote anatomical lobes. Pearson’s correlation analysis showed a significant negative association, indicating that higher GM proportion was associated with lower FD.

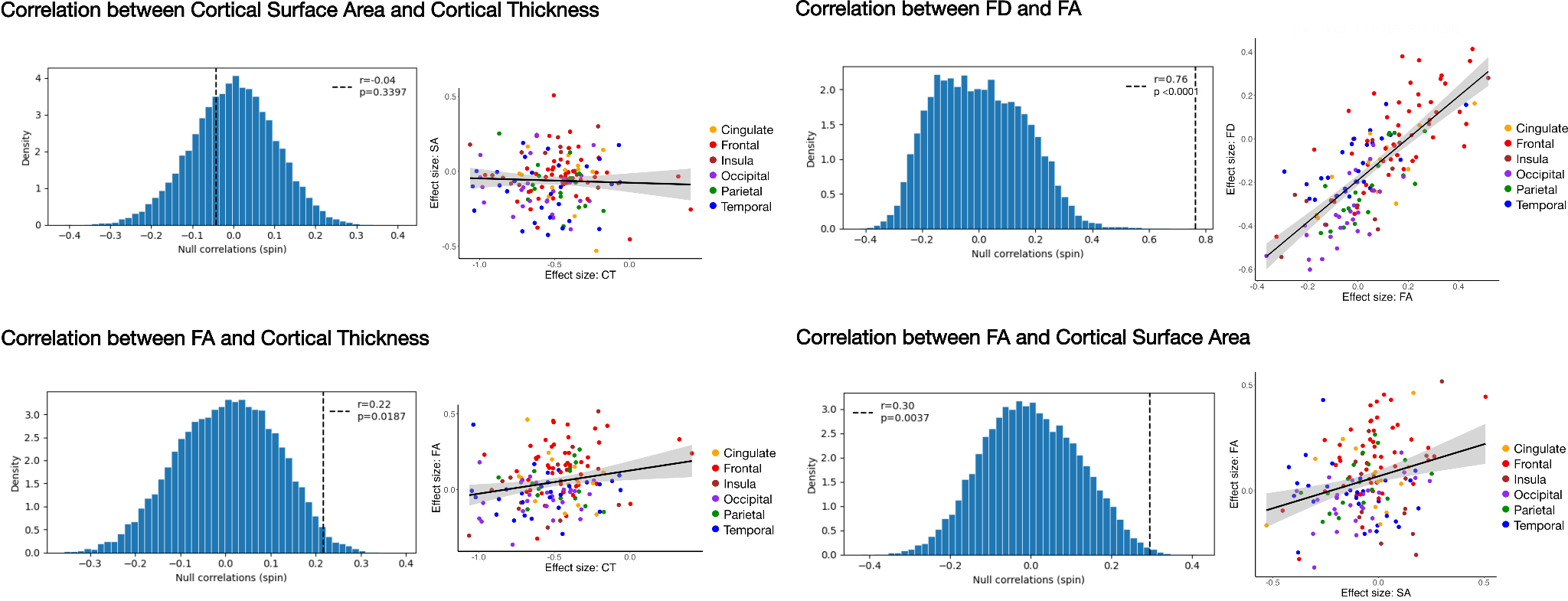

Figure S6. Spatial correlations between cortical surface area and thickness, between FD and FA, and between FA and cortical measures. Spatial correlations between regional effect sizes are shown. Upper left: cortical surface area vs. cortical thickness. Upper right: FD vs. FA. Lower left: FA vs. cortical thickness. Lower right: FA vs. cortical surface area. The left panels display the null distributions of spatial correlations generated using a spin test, with the vertical dashed line indicating the observed correlation. The right panels show scatter plots of the observed regional effect sizes with a fitted regression line. Each point represents a brain region, and colors indicate anatomical regions.

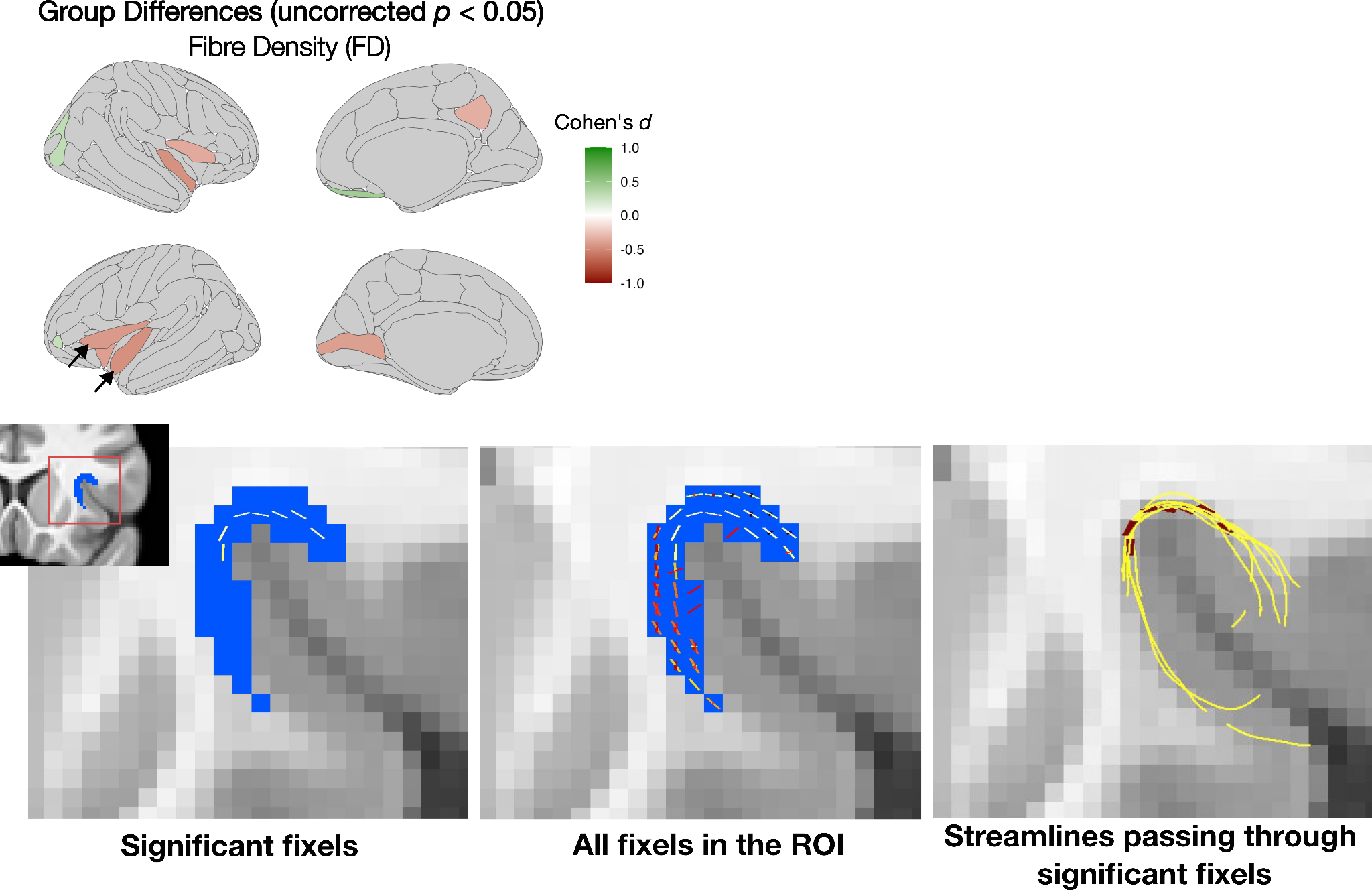

Figure S7. Exploratory fixel-based analysis in regions showing uncorrected significance.

The top panel shows significant regions at an uncorrected threshold in the template-based analysis. Fixel-based analysis was performed in the two regions indicated by arrows. The lower left panel shows fixels within the ROI that exhibited significantly reduced FD, and the lower middle panel shows all fixels. The lower right panel shows streamlines passing through significant fixels, illustrating that FD reductions were specifically localized to superficial white matter tracts connecting the inferior frontal gyrus and the insular region.

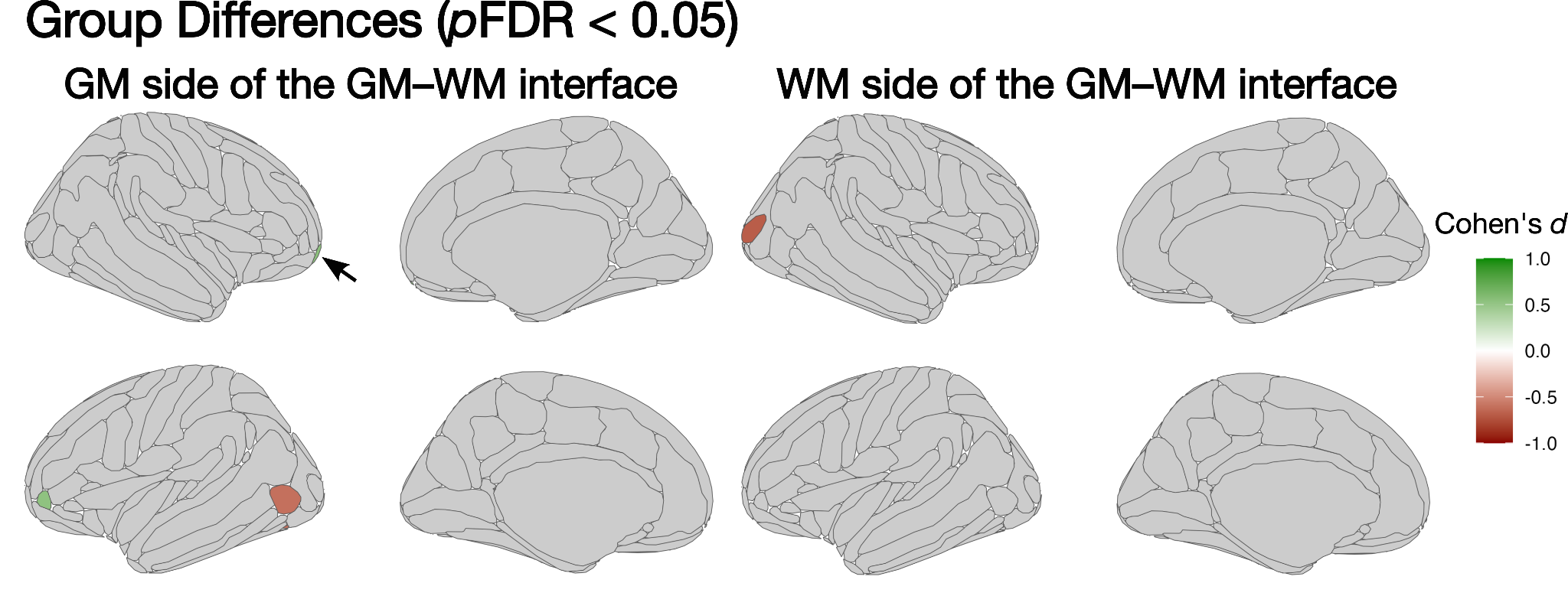

Figure S8. Group differences in FD of the GM and WM side of the GM–WM interface. Group differences in FD were assessed using regression models with age, sex, head motion, and GM proportion as covariates. Brain regions showed significant differences in FD between individuals with early psychosis and healthy controls (*p*FDR < 0.05). The cursor indicates the right frontomarginal gyrus and sulcus.
